# Landscape of maternal and neonatal care provision in Lubumbashi, Democratic Republic of Congo: Results from a 2023 health facility census

**DOI:** 10.1101/2025.07.04.25330874

**Authors:** Angèle Musau Nkola, Manuela Straneo, Aline Semaan, Peter M Macharia, Françoise Malonga Kaj, Alain Ngongo Ngashi, Nathalie Mulongo, Wivine Ngandu Malu, Joseph Debaif Mutombo Kayembe, Niclette Lakula, Abdulu Mahuridi, Tabitha Ilunga Mpoyi, Janvier Kubuya Bonane, Albert Tambwe-A-Nkoy Mwembo, Rehema Ouko, Lorenzo Libertini, Marie Alice Mosuse, Lenka Beňová, Abel Ntambue Mukengeshayi

## Abstract

Sixty million births are expected in urban sub-Saharan Africa in 2030. To address maternal and perinatal mortality, reliable data on facilities providing urban childbirth care is essential.

We describe a census of all health facilities in Lubumbashi, the second largest conurbation in the Democratic Republic of Congo, where such information was outdated. We systematically identified existing health facilities and collected data using a questionnaire on childbirth care provision and use. We analysed facility distribution across the 11 health zones, their level, capability and birth volumes.

Out of 1,455 facilities identified, 971 reported providing childbirth care. Facility numbers exceeded the list available to health authorities and were five-fold greater compared to a previous census in 2006. There was a predominance of the private sector (97% of facilities). Private facilities were mainly primary level with very low birth volumes. One third of facilities reported performing caesarean sections, and they were not limited to hospitals.

A dense network of primary level, low volume, for-profit facilities provided most childbirth care in Lubumbashi. Regulation of the private sector is necessary to improve quality and financial accessibility of childbirth care, in view of the planned rollout of the free childbirth care program in all provinces.

**Biographical note:** The authors of this study work together under the UrbanBirth Collective umbrella to examine maternal health in cities. We are a group of academic researchers based in the DRC and Belgium and provincial health decision-makers. Jointly, we developed this study, collected the data, analysed the results and presented them to the stakeholders in Lubumbashi. The main concern bring us together is the health of women, their babies and families in large cities in Africa, and how health systems can be designed or adapted to better meet their needs.

## Background

Maternal mortality remains a public health problem worldwide, with the highest burden in low-resource countries. Globally, around 260,000 women died from preventable causes relating to pregnancy and childbirth in 2023. The majority of deaths were concentrated in sub-Saharan Africa (SSA), with an estimated 182,000 deaths per year, representing 70% of maternal deaths worldwide (1).

The highest risk of maternal and perinatal death exists between the onset of labour until after childbirth (2). Most deaths can be prevented if women give birth in a health facility with the appropriate level of care to manage complications or with functional referral systems to reach appropriate health services in time, if obstetric complications occur. Thus, reducing maternal and perinatal mortality requires addressing inequalities in access to quality health services, and strengthening the health system (3).

Childbirth care in large cities in SSA presents unique challenges. According to United Nations’ estimates, by 2050, over two-thirds of global population will live in urban areas. In SSA, over 60 million births are expected to occur in urban settings by 2030. Rapid urbanization has placed significant pressure on health systems with limited capacity to adapt (4, 5). Partly in response to overcrowded, underfunded public facilities, a diverse and often poorly regulated private for-profit sector has emerged, contributing to poor quality of care (6). Evidence suggests that the “urban advantage” - historically linked to greater access to care and other infrastructure in urban settings - is stalling and possibly reversing (7, 8).

Efforts to improve maternal care in urban areas are hindered by the lack of comparable and reliable data on the complex dynamics of provision and utilization of childbirth care (9). In many cities, gaps in accurate, geocoded data on existing healthcare facilities make it challenging to develop effective strategies to improve maternal and perinatal health outcomes (10). In Lubumbashi, Democratic Republic of Congo (DRC), underfunding of the public sector has contributed to the unregulated expansion of the private sector for childbirth services (6). However, detailed information on health facilities in the city relies on outdated data, with the last comprehensive census conducted in 2006 (11).

The objective of this study was to examine the availability and functioning of health facilities providing childbirth care services in Lubumbashi city, DRC. We collected primary data in the form of a comprehensive census of all facilities. We accurately map their distribution, assess their provision of maternal and neonatal care services, and provide policymakers with a detailed landscape of childbirth care in the city.

## Methods

The protocol for this study was published (12). This study had an observational, cross-sectional design.

### Study setting

The study setting was Lubumbashi, DRC. In the country, maternal mortality ratio was one of the highest in the world in 2023 with 427 deaths per 100,000 live births, despite a reduction of 39% in the last thirty years (879 per 100,000 live births in 1990) (1). Key population and maternal and perinatal indicators for DRC and the West and Central African region are reported in **Table 1**. In DRC, healthcare is offered according to an integrated model based on a district health system. Maternal and perinatal care is organized in two tiers: the first line consists of health centres offering basic emergency obstetric and neonatal care (BEmONC), and in the second tier, general referral hospitals are authorized to provide comprehensive emergency obstetric and neonatal care (CEmONC) (13). In urban settings, women have direct access to the second line of care (i.e., hospitals) and choice of a referral facility within a context of a pluralist health system (public, private for-profit, private faith-based, private non-governmental) (14–16).

**Table 1.** Summary of maternal and neonatal indicators in the Democratic Republic of Congo and West & Central Africa region.

| Indicator | DR Congo | West & Central Africa |
| --- | --- | --- |
| Population (2023)(17) | 105,789,731 | 615,419,191 |
| Live births (2023) (17) | 4,369,683 | 21,418,323 |
| Urban population (% of total population) (2018) (18) | 45.6 | NA |
| Maternal mortality ratio (per 100,000 live births) (2023) (1) | 427 | 629 |
| Neonatal mortality rate (per 1,000 live births) (2023) (17) | 25.4 | 29.3 |
| Stillbirth rate (per 1,000 total births) (2023) (17) | 25.9 | 23.1 |

Lubumbashi is the second largest city in the DRC and the economic and social capital of the Haut Katanga province. In 2020, the city spanned an area of 870 km^2^; its population was estimated at 3.1 million in 2025 (18). In the year of the data collection (2023), approximately 121,520 births took place in Lubumbashi (based on a crude birth rate of 4.34%)(19). The city is subdivided into 11 urban health zones (HZ), among which two special, geographically small zones (Vangu - army and Kowe - police) are embedded in other health zones. Each HZ has one public General Referral Hospital (GRH). In addition to health facilities within HZs, there is a provincial referral hospital (Jason Sendwe) and the University Clinics of Lubumbashi (administered by the University of Lubumbashi). Health zones are further subdivided into health areas. Haut Katanga province had a very high percentage of births occurring in health facilities - 94% at the most recent population-representative survey (9).

A census of health facilities in Lubumbashi conducted in 2006 reported 251 facilities and their concentration in the city centre (core urban) (20). The study noted that the public sector accounted for less than 10% of these facilities. No other census of healthcare facilities or a detailed description of the provision of maternal/neonatal care had been carried out since. The health authorities in the 11 HZs did not have accurate and updated lists of all health facilities showing services offered at the time our study was conducted.

### Sampling

This was a census of all health facilities in Lubumbashi, focusing on those providing childbirth care. We first obtained a list of healthcare facilities from the Provincial Health Division (PHD) and supplemented it with lists provided by representatives of each HZ. This resulted in a total of 1,069 facilities of which an estimated 750 to 800 offered childbirth care. To ensure we identified all facilities, we included a snowballing technique to identify additional facilities by asking facility-based respondents to name the three closest healthcare facilities by perceived distance. All newly identified facilities were visited by the research team. Global positioning system coordinated GPS coordinates from every health facility in the city were taken during data collection, even those closed at the time of data collection.

### Data collection

#### Questionnaire and respondents

The questionnaire consisted of three sections. Respondents to the first part (Section 1) were selected based on their familiarity with the infrastructure, resources, and capacity of the health facility in general. When the facility was a hospital, this respondent was either the person in charge of reception or the manager. In lower-level facilities (health centers or health posts), the nurse in-charge (*infirmiere titulaire*) responded. If the facility provided childbirth care, questions from Section 2 focusing on childbirth care were asked (Figure 2) of a respondent with a thorough knowledge of the capacity, resources, and infrastructure of the maternity ward. Respondents to this section were either heads of the maternity ward, the midwife, maternity nurse or matron. Routine data from maternity ward registers on process indicators (number of births, caesarean (C-sections) and outcomes (live births, stillbirths, maternal complications) were collected from facilities which reported a birth volume of >30 births/month, or at least one caesarean section in the month before the survey (Section 3). Monthly aggregate routine data over a 12-month period (November 2022-October 2023) was extracted. Variables collected are summarized in Table 2. The full questionnaire is provided in the published protocol (12).

**Figure 1:**
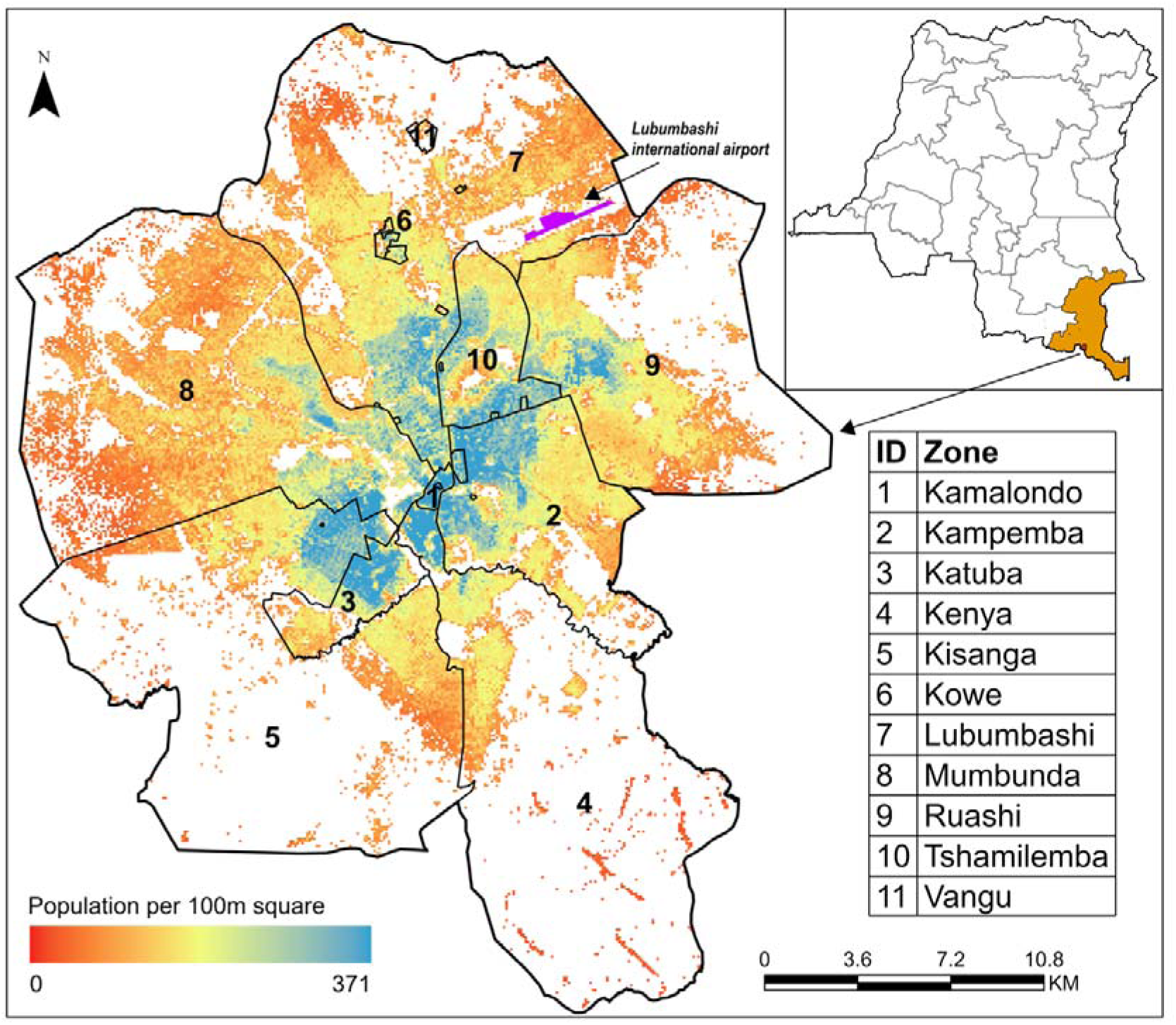
A map of Lubumbashi (study area), showing its subdivisions (health zones numbered 1 to 11) overlaid on its the population distribution in 2020 at 100m spatial resolution. The top right corner shows the location of Lubumbashi (red dot) within Haut Katanga province (orange) in the Democratic Republic of Congo.

**Figure 2.**
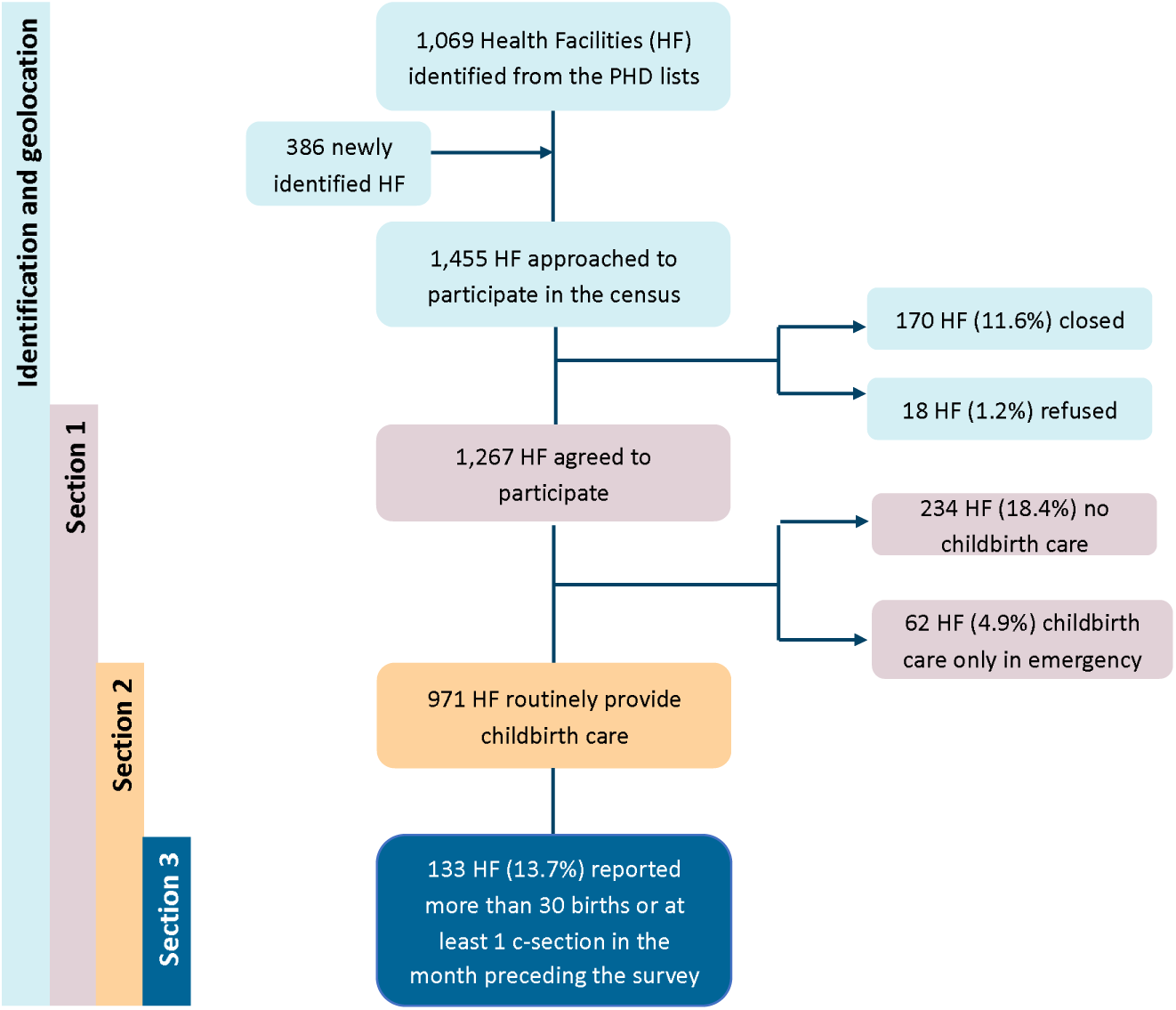
Flowchart of identification and inclusion of health facilities in a census, Lubumbashi DRC. HF health facilities, PHD Provincial Health Division, HZ Health Zone

**Table 2.** Description of variables analysed.

| Data source | Variable | Categorisation |
| --- | --- | --- |
| Respondent (Section 1) | Facility level | Eleven types of health facilities were categorized into three levels:<br>- **Primary** (*centre médical, centre de santé, centre de santé de référence, poste de santé*);<br>- **Secondary** (*hôpital, hôpital général de référence, centre hospitalier, clinique privée, polyclinique privée*);<br>- **Tertiary** (*hôpital provincial, cliniques universitaires*) |
|  | Facility sector | Facility ownership was grouped into sectors:<br>- **Public** (owned by the state);<br>- **Private for-profit** (owned by an individual, a group of individuals, or a private organization);<br>- **Private non-profit** (*paraétatique* or owned by a public organization, a non-governmental organization, a confessional organization, or a church/other religious institution). |
|  | Opening hours | The hours during which the health facility is open to patients for consultations or care by health personnel (in two categories: open 24/7, open less than 24/7). |
|  | SNIS integration | Facility integration into the *Système National d'Information Sanitaire* (National Health Information System) which is a DHIS2-based process for organizing information and producing the indicators needed for health system management, monitoring, evaluation, and planning health interventions. Response: Yes; No. |
|  | Financial support from partners | Whether the facility received financial support from partners for its operation. Response: Yes; No. |
| Respondent (Section 2) | Emergency obstetric care | This was assessed using the Emergency Obstetric and Neonatal Care framework (23). The following nine signal functions were used: 1) administration of intravenous antibiotics, 2) administration of IV/IM oxytocics, 3) administration of parenteral magnesium sulphate, 4) assisted vaginal birth, 5) manual extraction of placenta, 6) removal of retained products of conception, 7) neonatal resuscitation with bag and mask, 8) cesarean section, 9) blood transfusion. Due to expected low volumes of births in facilities, we adapted the timeframe of the indicators to the previous 12 months, as the standard definition (functions must have been performed at least once in the previous three months) may not fully capture obstetric capability in this setting. The first seven indicators were used for BEmONC, while all nine represented CEmONC. Due to limited use of assisted vaginal delivery in this context, we additionally combined signal functions in two modified indicators, BEmONC-1 (all BEmONC functions except assisted vaginal birth, total six functions) and CEmONC-1 (all CEmONC functions except assisted vaginal birth, total eight functions), as in other studies (24). Other facilities were classified as non-EmONC. |
|  | Reported facility birth volume | The number of births in the facility in the month before the survey, as reported by the respondent (in four categories, <10 births/month, 10-30 births/month, >30 births/month, unknown). |
| Maternity ward registers (Section 3) | Births | Number of births in the facility in a given calendar month. |
|  | C-section rate | Percentage of births via C-section from the total number of births in the facility. |
|  | Stillbirth rate | Rate of stillbirths per 1,000 births in the facility. |

#### Data collection

A team of 25 trained data collectors used tablets to collect the data with KoboCollect application (21) between November and December 2023. When no respondent was available to answer the questionnaire, a follow-up visit was planned with a scheduled appointment. A team leader/supervisor for each group of 8 to 10 data collectors supervised the field activities. Missing or inconsistent data were retrieved and/or corrected in January 2024.

#### Data quality checks

Strategies to avoid unlikely and inconsistent responses were pre-coded in the digital data collection forms. These included embedding automatic skip-patterns for each section of the questionnaire to reduce errors in the identification of the subsets of healthcare facilities. Collected data was reviewed daily by field supervisors for quality assurance. We also performed additional data quality checks during the fieldwork, which allowed the interviewers to return to health facilities or call respondents if there were data quality issues. This included, for example, identifying facilities that were interviewed twice by different enumerators or two facilities with the same global position system coordinates coordinates.

### Data analysis

Preliminary results of the census were presented to the PHD and representatives of all 11 HZs in the form of maps and tables in July 2024 and October 2024 (22). Their questions and priority needs were reflected in the objective of this paper. Analysis was performed using STATA SE16. We described baseline characteristics of all facilities and of those providing childbirth care, using percentages. We further analysed numbers and percentage of facilities providing childbirth care by sector and level, and by birth volumes. The distribution of facilities per HZ was examined. We examined availability of obstetric functions using the signal functions’ framework (23). In a subset of facilities with ≥30 births/month or at least one C-section (eligible for questionnaire Section 3), we analysed data extracted from maternity ward registers. To estimate yearly totals of facility births, C-sections, and stillbirths, where data were missing, we extrapolated using the average of months available. We examined process and outcome indicators in facilities using the internationally recognized threshold of 500 births/year.

### Ethical considerations

The study was approved by the Institute of Tropical Medicine’s Institutional Review Board, ITM (1715/23) and the Medical Ethics Committee of the University of Lubumbashi (UNILU/CEM/028/2023). Data collectors and researchers received written permission from health zone officials to visit the healthcare facilities. Informed consent to participate in this study was obtained from all facility-based respondents. Participants who had agreed to participate in the study were asked to put a digital signature on the tablet to attest to their written informed consent.

## Results

### Characteristics of all health facilities

We identified and approached 1,455 healthcare facilities (1,069 facilities from health authorities’ lists and 386 additional facilities identified through snowballing). Of the total, 170 facilities were closed and representatives of 18 refused to participate in the census, leading to a total of 1,267 facilities in which data collection was successfully conducted. Childbirth care was routinely provided by 971 of these facilities. We extracted routine data from a subset of 133 of facilities reporting ≥30 births or at least one C-section in the month preceding the survey (Figure 2). Basic characteristics of facilities surveyed and of those which provided childbirth care are shown in Table 3.

**Table 3.** Characteristics of facilities included and subset of facilities providing childbirth care, Lubumbashi DRC.

| Variable | Category | All facilities<br>n, (%) | Facilities providing<br>childbirth care n, (%) |
| --- | --- | --- | --- |
| N |  | 1,267 | 971 |
| Health zone | Kamalondo | 17 (1.3%) | 10 (1.0%) |
|  | Kampemba | 243 (19.2%) | 190 (19.6%) |
|  | Katuba | 50 (3.9%) | 36 (3.7%) |
|  | Kenya | 126 (9.9%) | 78 (8.0%) |
|  | Kisanga | 128 (10.1%) | 94 (9.7%) |
|  | Kowe | 6 (0.5%) | 5 (0.5%) |
|  | Lubumbashi | 225 (17.8%) | 172 (17.7%) |
|  | Mumbunda | 157 (12.4%) | 130 (13.4%) |
|  | Ruashi | 222 (17.5%) | 181 (18.6%) |
|  | Tshamilemba | 88 (6.9%) | 71 (7.3%) |
|  | Vangu | 5 (0.4%) | 4 (0.4%) |
| Sector | Public | 43 (3.4%) | 31 (3.2%) |
|  | Private for-profit | 1,087 (85.8%) | 825 (85.0%) |
|  | Private non-profit | 137 (10.8%) | 115 (11.8%) |
| Level | Primary | 1,128 (89.0%) | 853 (87.8%) |
|  | Secondary | 137 (10.8%) | 116 (11.9%) |
|  | Tertiary | 2 (0.2%) | 2 (0.2%) |
| SNIS integration | Yes | 1,089 (86.0%) | 893 (92.0%) |
|  | No | 170 (13.4%) | 76 (7.8%) |
|  | Not known | 8 (0.6%) | 2 (0.2%) |
| Financial support from partners | Yes | 32 (3.3%) | 32 (3.3%) |
|  | No/not known | 939 (96.7%) | 939 (96.7%) |
| Facility opening hours 24/7 | Yes | 1,208 (95.3%) | 958 (98.7%) |

### Characteristics of facilities providing childbirth care services

The majority of the 971 facilities providing childbirth care were private (96.8%) - private for-profit were 825 (85.0%) and 115 (11.8%) were non-profit (**Supplementary Table 1**). Among private for-profit facilities, 88.8% were primary facilities (733), and 11.2% were secondary facilities. There were no tertiary facilities in the private for-profit sector. In the private non-profit sector, the distribution was similar, with 98 (85.2%) of primary, 16 (13.9%) secondary, and one tertiary facility. In the public sector, there were 22 (71.0%) primary facilities, 8 (25.8%) secondary facilities, and one tertiary hospital. Over 90% of facilities in each sector reported integration into the SNIS routine data. Nearly all facilities were open 24 hours every day (24/7): only 2/31 (6.5%) public facilities and 11/825 (1.3%) private for-profit reported shorter opening hours. Most facilities which reported providing childbirth services were primary facilities, 853/971 (87.8%). There was variability across the HZs in the distribution of facilities. Lubumbashi HZ had both (2/2) tertiary hospitals. This zone also had the majority of secondary (34/116, 29.3% of secondary facilities) and 15.9% of primary facilities (136/853) (**Supplementary Table 2**). The lowest number of facilities was in the two special HZs, Kowe and Vangu.

### Emergency obstetric care functions

We found that the most commonly reported obstetric signal functions performed in the previous 12 months were administration of IV antibiotics and IV/IM oxytocics, in over 90% of facilities across all sectors. Assisted vaginal birth was the least commonly reported function, in 30-35% of facilities in all sectors (**Supplementary Table 3 and Supplementary Figure 1**). Using the EmONC framework, around half of facilities in all sectors were non-EmONC (500/825, 61% in private for-profit). Between 6-15% of facilities in all sectors reported all BEmONC functions in the previous 12 months. An additional 6-10% were classified as BEmONC-1 (without assisted vaginal birth) (**Figure 3-A**).

**Figure 3.**
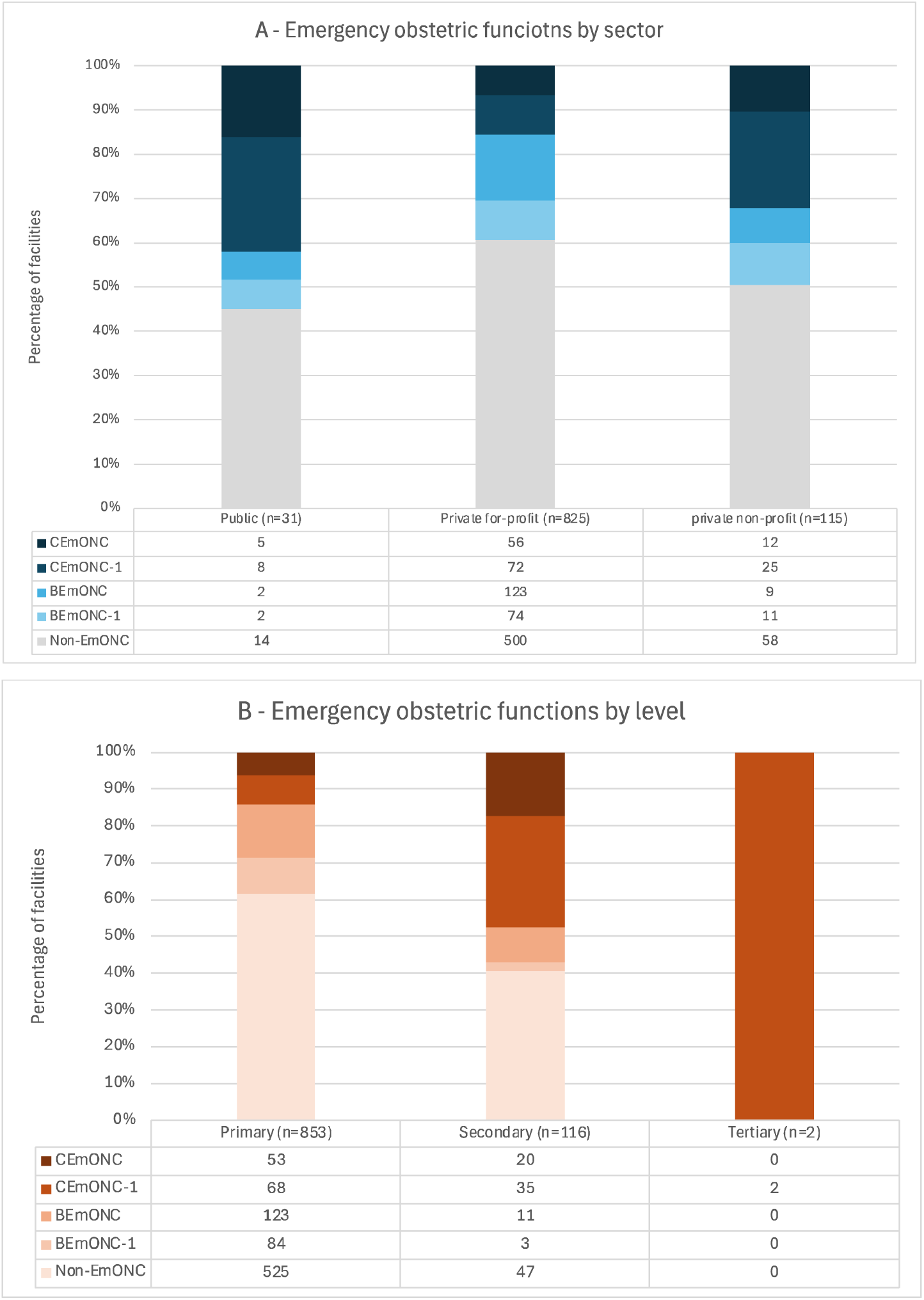
Emergency obstetric care functions performed in the previous 12 months in Lubumbashi (n=971, facilities providing childbirth care) **A. By sector** **B. By level** BEmONC - Basic emergency and neonatal care functions (7 functions) BEmONC-1 Basic emergency and neonatal care functions except assisted vaginal delivery (6 functions) CEmONC Comprehensive emergency and neonatal care functions (9 functions) CEmONC-1 Comprehensive emergency and neonatal care functions except assisted vaginal delivery (8 functions) Non-EmONC Facilities that could not be classified in any of the above, meaning they have fewer than BEmONC-1 functions

Functions assessing more advanced management of childbirth complications (C-section and blood transfusions) were most common in the public sector and least in the private for-profit sector. A total of 289 facilities reported having performed a C-section in the previous 12 months. C-section was performed in 22/31 (71.0%) of public facilities and in 208/825 (25.2%) of private for-profit facilities. Blood transfusions were performed in 27/31 (87.1%) public facilities, and in 492/825 (59.6%) private for-profit facilities. CEmONC availability was 7-16% in all sectors (highest in the public sector), with an additional 9-26% of facilities classified as CEmONC-1 (without assisted vaginal birth). (**Figure 3-A**).

The percentage of facilities that had performed signal functions increased with level (**Figure 3B**, **Supplementary Table 4)**. C-sections and blood transfusions were more common in tertiary than in secondary facilities. C-sections were reported in 73.3% of secondary facilities, and in both (100%) tertiary facilities. Similarly, blood transfusions were reported 83.6% of secondary facilities and 100% of tertiary facilities. Due to no reported assisted vaginal births, neither tertiary facility could be classified as providing comprehensive EmONC, but both fulfilled the CEmONC-1criteria (**Figure 3-B**).

### Emergency obstetric care functions in primary facilities

The majority (62%) of primary facilities were non-EmONC; 14% reported performing all BEmONC functions in the previous 12 months and BEmONC-1 was reported in an additional 10% of facilities. C-sections were reported in 202/853 (23.7%) of primary care facilities. The percentage of primary facilities reporting C-section provision was highest in public sector 13/22 (59.1%) and lowest in the private for-profit (147/733, 20.1%). Blood transfusions were reported in 509/853 (59.7%) of primary facilities, more commonly in public ones (18/22, 81.8%) and least in private for-profit (419/733, 57.2%). We found that 53/853 primary facilities reported all CEmONC functions. These were more common in public facilities (18.3%) and least in private for-profit facilities (5.6%). When we excluded assisted vaginal birth (CEmONC-1), 121/853 facilities reported all eight CEmONC functions; ranging from 31.8% of public facilities to 12.3% of private for-profit facilities (12.3%).

### Facility birth volumes

Birth volumes varied by health zone. Kampemba and Mumbunda HZs had the highest percentage of low volume facilities (over half of facilities providing childbirth care reported <10 births/month), and Kisanga the lowest (33.0%), **Supplementary table 5**. Reported volumes of births per month varied by sector and level. In the public sector, the majority of facilities reported 10-30 births/month (23/31, 74.2%), and 5/31 (16.1%) reported >30 births/month. In the private for-profit sector, 419/825 (50.8%) facilities reported under 10 births/month. In the private non-profit sector, there was a higher percentage of facilities reporting >30 births monthly (26/115, 22.6%). Both tertiary hospitals reported volumes above 30 births/month. Among secondary facilities, the majority 63/115 (53.4%) reported birth volumes between 10-30 births/month. Among primary facilities, nearly half 423/853 (49.6%) reported less than 10 births/month.

### Analysis of routine data in facilities providing childbirth care (subset of facilities)

Among the 971 facilities with childbirth care, 133 facilities reported attending ≥30 births or having performed at least one C-section in the month before data collection and were therefore asked to provide monthly routine maternity ward data for the period November 2022 to October 2023. Analysis of the extracted routine data showed that the annual number of births among this subset of facilities ranged from 8 to 904, with a median of 298 in the 12-month period. However, these data had a high degree of missingness. Ten of the 133 facilities had ≥ 50% missing months, and for one facility, all records were missing due to the loss of paper-based maternity records (**Supplementary table 6**). A total of 102 facilities reported at least one C-section in the 12-month period (range 1 to 199), with a median of 11. For the main complications and adverse outcomes examined (eclampsia, antepartum hemorrhage, stillbirths), over half facilities had ≥50% missing months in the year examined, and around one quarter had all months missing.

After adjusting for missing months in the 132 facilities where this was possible, 100 facilities (78.8%) had birth volumes ≤500 births/year, 31 had a volume 501-1,000/year, and one facility had ≥1,000 births per year. In 102 facilities which reported at least one C-section, following adjustment for missing months, the percentage of C-sections out of total births ranged from 0.5% to 87%; 59% of facilities reported C-section rate of ≤10%, 23.5% reported 10.1-20% C-section rate, and 17.6% reported >20%.

Characteristics and outcomes in facilities were stratified by birth volumes (**Table 4**). Among lower birth volume facilities (≤500 yearly births), 11.0% belonged to the public sector, 69.0% to the private for-profit and 20.0% to the private non-profit sector. Among facilities with higher birth volumes (>500 births per year), 6.3% were public, 43.8% were private for-profit and 50% belonged to the private non-profit sector. By level, lower-volume facilities were primary (58.0%) or secondary level (42%). Higher-volume facilities (>500 births/year) were mostly primary (78.1%), with 15.6% secondary and 6.3% were tertiary. The majority of health facilities reporting C-sections were lower volume health facilities; 20% of them reported a C-section rate over 20%. Among higher volume facilities, only 12% reported a C-section rate over 20%. Higher rates of stillbirths (>100 per 1,000 births) were similar in lower and higher-volume facilities.

**Table 4.**
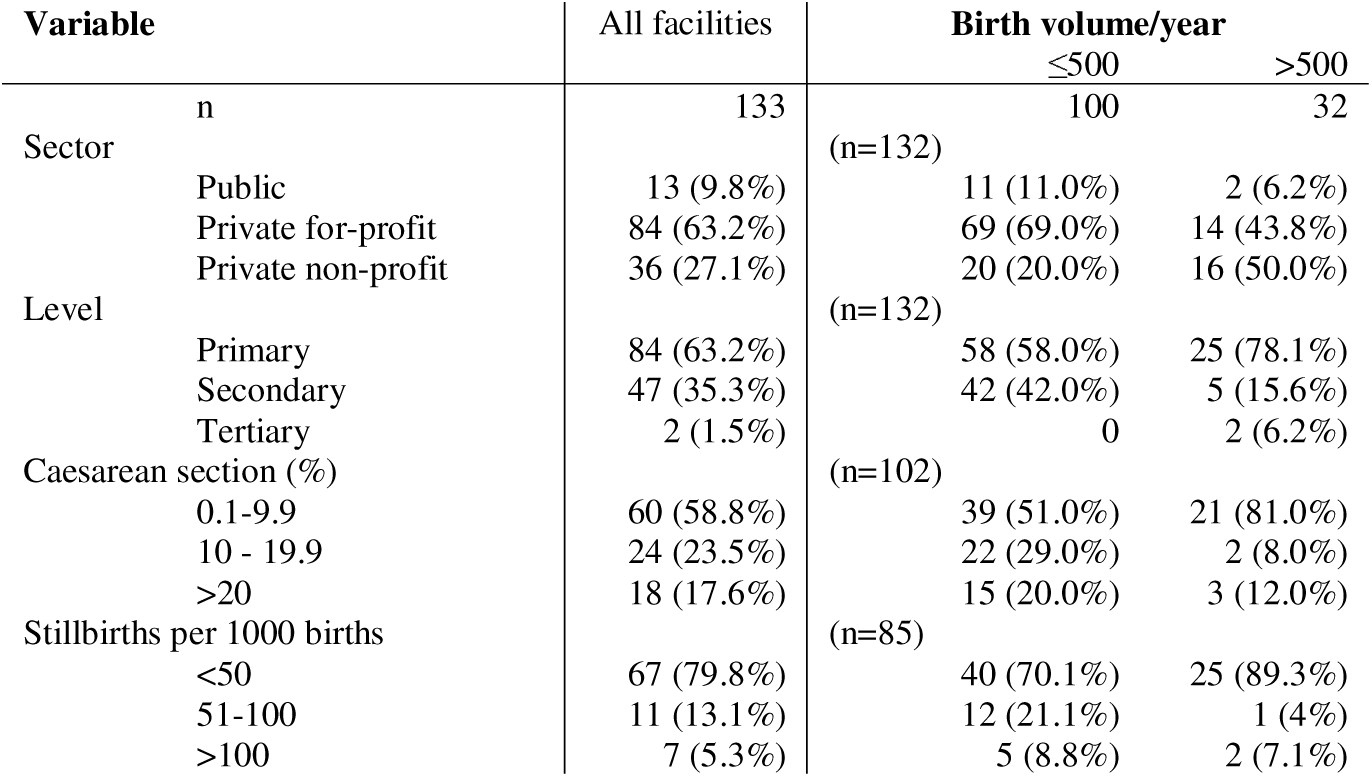
Key process and outcome indicators in facilities with at least 30 births or one C-section reported in the month before the census (n=133), by annual birth volume.

## Discussion

This study describes the characteristics of 971 facilities providing childbirth care identified through a comprehensive census in Lubumbashi, a large conurbation in the DRC. We highlight four main findings. First, there was a substantial growth in the number of health facilities since the previous census in 2006. Second, we found a near predominance of the private sector in provision of childbirth care. Third, approximately one third of facilities reported performing C-sections – including those at primary level; hospitals were not the majority providers of C-sections. Fourth, routine maternity ward data in facilities had high levels missingness and therefore limited usefulness for urban health system decision-makers.

The number of health facilities in Lubumbashi rose dramatically since the previous census 17 years ago (11). Our study identified 1,455 health care facilities, a six-fold increase since 2006. Childbirth care was available in two thirds of facilities (971), despite governmental guidelines that it should be available in all facilities (25). This corresponds to approximately 40 new facilities providing childbirth care being established *each year*, an average annual growth rate of 17% compared to the annual population growth rate of 4% (26). This exponential growth was predominantly driven by new primary facilities, a four-fold increase since 2006. The number of facilities exceeded those listed by local government authorities, suggesting that the full landscape of care provision is not entirely known to health authorities, adding challenges to regulation and monitoring in a rapidly urbanizing metropolis.

Second, we note the near dominance of the private sector in providing childbirth care in Lubumbashi, with 97% of facilities private, an increase from 75% in 2006 (20). Some of these facilities are currently outside the control of the local health authorities, resulting in a lack of information on service delivery, quality of care and poor integration in the SNIS (4). Further, our census showed that the private sector - both for-profit and non-profit – is constituted mainly of primary facilities (>85%). This near dominance of the private sector is not seen in other cities. In the Grand Conakry (Guinea Conakry) urban area, with a population of 2.6 milion in 2024, childbirth care was provided by 155 health facilities, one third belonging to the public sector (27). A higher percentage of private facilities (71% for-profit) was found in the Grand Nokouè metropolitan area (Benin) (28).

This provision landscape in Lubumbashi, with a high number of primary, private sector, low birth volume facilities providing the majority of facility-based childbirth care, is relatively recent. In the decade 2010-2019, high volumes in few facilities (9,000 births/year in one tertiary hospital) (6) were reported. At present, 48% of facilities attended under 10 births per month, and the highest volume facility reported just over 1,000 yearly births. The picture is different from other large urban conglomerates, where congestion of hospitals is considered a major barrier to quality of care, such as in Dar es Salaam, Tanzania (29), and in Kampala, Uganda (30). In these cities, facility birth volumes of >10,000 yearly are not uncommon. On the other hand, the increase of births in lower-level facilities has been described in other settings (24, 31), though these studies did not focus on urban areas.

Limited investment in the public sector since the colonial period can at least partly explain our findings. With the decline in the DRC’s economy during the 1990s, the country was unable to finance the healthcare system sufficiently to meet its needs or to operationalize the plan to extend healthcare coverage (32). Lack of regulation has resulted in the growth of private facilities - not in response to health needs, but for commercial purposes (6). Private facilities, operating in a competitive, market-like environment, often build strong relationships with users, developing trust with women and their families. In contrast, frequent strikes and shortages in the public sector undermine such trust. The near dominance of the private sector, together with very low health insurance coverage (33) also means that financial burden of chidbirth care in Lubumbashi is high, with cases of imprisonment for non-payment of fees being a common occurrence in both sectors (6).

Dense landscapes of competing private primary care clinics have been recently described across large cities in low-and middle-income countries (34). What our study adds is that the picture is similar for childbirth services in a large urban conurbation in DRC. We draw attention to the characteristics of childbirth care, which is different from other primary care services, due to the need to manage or refer unpredictable complications rapidly. Most facilities had low birth volumes: meaning that key skills are not practiced frequently. In high-resource settings, births have been progressively centralized to higher level, higher volume facilities to reduce morbidity and mortality during childbirth (35). Though there is no clear evidence on optimal birth volumes in primary care facilities in low-resource settings (36), low birth volumes are a challenge for staff skill retention, cost-effectiveness and, most importantly, for women and babies’ safety. The transition to lower maternal, perinatal mortality across countries is associated to a shift in births from lower level facilities to hospitals (37), suggesting that reducing births in low-volume facilities while ensuring good quality of care in higher-level facilities in this setting should be part of a strategy aiming to reduce maternal and perinatal mortality (38).

Third, C-sections were reported by a sizable percentage of facilities (30%). Hospitals were not the main providers of C-sections, as approximately one quarter (23.7%) of primary facilities reported providing C-sections. Analysis of maternity ward data showed that most facilities (58.8%) with at least one C-section in the month before the survey reported a low C-section rate (<10%), and volumes were low. In addition, C-sections were not limited to facilities with relatively higher volumes, highlighting the importance of emergency referral between facilities when complications arise. This distribution of C-sections is difficult to compare to other settings in SSA, as C-sections are generally limited to hospitals, which have higher C-section rates. For example, in a study across 16 hospitals in four countries in SSA, C-section rates ranged between 17.3-45.5% (39). The large number of facilities providing C-sections in Lubumbashi makes it difficult to supervise facilities and ensure quality of care. The private for-profit sector remains the predominant provider of C-sections, raising questions about quality of care, the financial accessibility and the risk of catastrophic expenses at the time of childbirth. Despite these high numbers of facilities offering C-sections, we might have underestimated the number because of fear of respondents to report the practice, especially in primary facilities.

The EmONC framework had to be adapted to this setting, using a time interval for signal functions of 12 months as opposed to 3 months (23). With this adaptation, the number of CEmONC facilities exceeded the WHO recommendation of one CEmONC per 500,000 population. We note that assisted vaginal birth (vacuum or forceps) was rarely reported. As a result, no tertiary facility could be classified as providing CEmONC, though some secondary (n=20) and primary (n=53) level facilities reported providing all functions. Low use of assisted vaginal birth has been reported in other settings (24), and may represent an opportunity to avoid second-stage C-sections and reduce maternal and perinatal mortality (40). Similarly to C-sections, blood transfusions were not limited to secondary or tertiary facilities, as they were performed in 60% of primary care facilities. Only 14% of facilities were classified as providing BEmONC, similarly to what reported in other parts of the country (25).

Lastly, routine maternity ward data (births, C-sections, stillbirths, and maternal complications) had high levels of missingness. For example, around one quarter of facilities performing C-sections had six or more months out of 12 with missing data. This questions the existing documentation system in health facilities and its usefulness for monitoring process and outcomes of childbirth care, as has been reported in other settings (41, 42). It is likely that these deficiencies are present in facilities’ reports to the DHIS2/SNIS and therefore have limited value to decision-makers. Limitations of these data should also be considered in view of the potential under-reporting of certain procedures such as C-sections performed at the primary care facility level. This finding underlines the need to strengthen routine data collection. The large number of health facilities, with a substantial private sector presence, make supportive supervision challenging (25).

### Strengths and limitations

The comprehensiveness of this facility census and focus on childbirth care is a major strength of this study. However, some limitations exist. Data may be subject to recall bias. For example, reporting of facility birth volumes may lack accuracy. Most importantly, under-reporting of signal functions (especially C-sections) and birth volumes might have occurred due to fear of closure or financial repercussions (e.g., taxes on income). Low numbers of C-sections may limit accuracy of reporting; for instance, 230 facilities in section 2 reported performing C-sections, though in section 3, 102 facilities reported at least one C-section in the previous month, with the difference likely due to not performing the procedure In the previous month. Annual birth volumes might have been underestimated due to seasonal variations.

The use of the signal function framework (23) has limitations, and we cautiously note that it may be unsuitable for this low-volume setting. Complications such as retained placenta and eclampia are relatively rare (<5% of births in population) (43, 44), thus staff in facilities may have the necessary training, but the complication may not be seen in the time frame captured on our questionnaire. However, these limitations do not prevent us from describing in detail the portrait of maternal and neonatal health care provision in accordance with the universal public health policy, and from making concrete recommendations for stakeholders at the city, provincial and national levels.

## Conclusion

This study reported a census of health facilities providing childbirth care in Lubumbashi. Facilities identified and described exceeded the list available to health authorities. The private for-profit sector was predominant, and facilities were mainly primary and low volume. C-sections were performed by around a third of facilities, and were not limited to hospitals. Regulation and monitoring of these large numbers of facilities is challenging but essential to reduce maternal and perinatal mortality.

## Supporting information

Manuscript

## Data Availability

All data produced in the present study are available upon reasonable request to the authors

## Acknowledgments

We acknowledge the essential support of the data collectors. We also thank respondents in health facilities, who gave their time to answer our questions, and health zone representatives who granted permission to conduct the filedwork and provided lists of health facilities.

