## Supplementary material for "Landscape of maternal and neonatal care provision in Lubumbashi, Democratic Republic of Congo: Results from a 2023 health facility census": Manuscript

**Supplementary Table 1. Characteristics of facilities providing childbirth care by sector, Lubumbashi (n=971)**

| Factor | Category | Public | Private for-profit | Private non-profit |
| --- | --- | --- | --- | --- |
| N |  | 31 | 825 | 115 |
| Level | Primary | 22 (71.0%) | 733 (88.8%) | 98 (85.2%) |
|  | Secondary | 8 (25.8%) | 92 (11.2%) | 16 (13.9%) |
|  | Tertiary | 1 (3.2%) | 0 (0.0%) | 1 (0.9%) |
| Health zone | Kamalondo | 1 (3.2%) | 7 (0.8%) | 2 (1.7%) |
|  | Kampemba | 3 (9.7%) | 166 (20.1%) | 21 (18.3%) |
|  | Katuba | 2 (6.5%) | 27 (3.3%) | 7 (6.1%) |
|  | Kenya | 5 (16.1%) | 66 (8.0%) | 7 (6.1%) |
|  | Kisanga | 2 (6.5%) | 85 (10.3%) | 7 (6.1%) |
|  | Kowe | 3 (9.7%) | 2 (0.2%) | 0 (0.0%) |
|  | Lubumbashi | 3 (9.7%) | 135 (16.4%) | 34 (29.6%) |
|  | Mumbunda | 4 (12.9%) | 114 (13.8%) | 12 (10.4%) |
|  | Ruashi | 2 (6.5%) | 163 (19.8%) | 16 (13.9%) |
|  | Tshamilemba | 2 (6.5%) | 60 (7.3%) | 9 (7.8%) |
|  | Vangu | 4 (12.9%) | 0 (0.0%) | 0 (0.0%) |
| SNIS integration | Yes | 30 (96.8%) | 753 (91.3%) | 110 (95.7%) |
|  | No | 1 (3.2%) | 70 (8.5%) | 5 (4.3%) |
|  | Unknown | 0 (0.0%) | 2 (0.2%) | 0 (0.0%) |
| Facility open 24/7 | No | 2 (6.5%) | 11 (1.3%) | 0 (0.0%) |
|  | Yes | 29 (93.5%) | 814 (98.7%) | 115 (100.0%) |
| Reported birth volume | <10 per month | 3 (9.7%) | 419 (50.8%) | 44 (38.3%) |
|  | 10-30 per month | 23 (74.2%) | 368 (44.6%) | 45 (39.1%) |
|  | >30 per month | 5 (16.1%) | 36 (4.4%) | 26 (22.6%) |
|  | Unknown | 0 (0.0%) | 2 (0.2%) | 0 (0.0%) |

**Supplementary Table 2. Characteristics of facilities providing childbirth care by level, Lubumbashi (n=971)**

| Factor | Category | Primary | Secondary | Tertiary |
| --- | --- | --- | --- | --- |
| N |  | 853 | 116 | 2 |
| Sector | Public | 22 (2.6%) | 8 (6.9%) | 1 (50.0%) |
|  | Private for-profit | 733 (85.9%) | 92 (79.3%) | 0 (0.0%) |
|  | Private non-profit | 98 (11.5%) | 16 (13.8%) | 1 (50.0%) |
| Facility open 24/7 |  | 841 (98.6%) | 115 (99.1%) | 2 (100.0%) |
| Health zone | Kamalondo | 8 (0.9%) | 2 (1.7%) | 0 (0.0%) |
|  | Kampemba | 173 (20.3%) | 17 (14.7%) | 0 (0.0%) |
|  | Katuba | 33 (3.9%) | 3 (2.6%) | 0 (0.0%) |
|  | Kenya | 76 (8.9%) | 2 (1.7%) | 0 (0.0%) |
|  | Kisanga | 86 (10.1%) | 8 (6.9%) | 0 (0.0%) |
|  | Kowe | 5 (0.6%) | 0 (0.0%) | 0 (0.0%) |
|  | Lubumbashi | 136 (15.9%) | 34 (29.3%) | 2 (100.0%) |
|  | Mumbunda | 109 (12.8%) | 21 (18.1%) | 0 (0.0%) |
|  | Ruashi | 166 (19.5%) | 15 (12.9%) | 0 (0.0%) |
|  | Tshamilemba | 59 (6.9%) | 12 (10.3%) | 0 (0.0%) |
|  | Vangu | 2 (0.2%) | 2 (1.7%) | 0 (0.0%) |
| Reported birth volume | <10 per month | 423 (49.6%) | 43 (37.1%) | 0 (0.0%) |
|  | 10-30 per month | 374 (43.8%) | 62 (53.4%) | 0 (0.0%) |
|  | >30 per month | 54 (6.3%) | 11 (9.5%) | 2 (100.0%) |
|  | unknown | 2 (0.2%) | 0 (0.0%) | 0 (0.0%) |

**Supplementary Table 3.** **Reported performance of emergency obstetric care functions (in the previous 12 months), by sector (among facilities providing childbirth care n= 971)**

| Obstetric function | Category | Public | Private for-profit | Private non-profit | All facilities |
| --- | --- | --- | --- | --- | --- |
| N |  | 31 | 825 | 115 | 971 |
| Administration of IV antibiotics | yes | 30 (96.8%) | 782 (94.8%) | 110 (95.7%) | 922 (95.0%) |
|  | no, but facility has capability | 1 (3.2%) | 24 (2.9%) | 3 (2.6%) | 28 (2.9%) |
|  | no/unknown | 0 (0.0%) | 19 (2.3%) | 2 (1.7%) | 21 (2.2%) |
| Administration of IV/IM oxytocics for PPH | yes | 29 (93.5%) | 765 (92.7%) | 111 (96.5%) | 905 (93.2%) |
|  | no, but facility has capability | 2 (6.5%) | 38 (4.6%) | 3 (2.6%) | 43 (4.4%) |
|  | no/unknown | 0 (0.0%) | 22 (2.7%) | 1 (0.9%) | 23 (2.4%) |
| Administration of parenteral magnesium sulphate for eclampsia | yes | 27 (87.1%) | 601 (72.8%) | 96 (83.5%) | 724 (74.6%) |
|  | no, but facility has capability | 4 (12.9%) | 125 (15.2%) | 16 (13.9%) | 145 (14.9%) |
|  | no/unknown | 0 (0.0%) | 99 (12.0%) | 3 (2.6%) | 102 (10.5%) |
| Assisted vaginal birth | yes | 11 (35.5%) | 278 (33.7%) | 35 (30.4%) | 324 (33.4%) |
|  | no, but facility has capability | 3 (9.7%) | 68 (8.2%) | 15 (13.0%) | 86 (8.9%) |
|  | no/unknown | 17 (54.8%) | 479 (58.1%) | 65 (56.5%) | 561 (57.8%) |
| Manual extraction of placenta | yes | 24 (77.4%) | 713 (86.4%) | 105 (91.3%) | 842 (86.7%) |
|  | no, but facility has capability | 5 (16.1%) | 83 (10.1%) | 8 (7.0%) | 96 (9.9%) |
|  | no/unknown | 2 (6.5%) | 29 (3.5%) | 2 (1.7%) | 33 (3.4%) |
| Removal of retained products of conception | yes | 31 (100.0%) | 750 (90.9%) | 106 (92.2%) | 887 (91.3%) |
|  | no, but facility has capability | 0 (0.0%) | 54 (6.5%) | 4 (3.5%) | 58 (6.0%) |
|  | no/unknown | 0 (0.0%) | 21 (2.5%) | 5 (4.3%) | 26 (2.7%) |
| Neonatal resuscitation with bag and mask | yes | 23 (74.2%) | 424 (51.4%) | 75 (65.2%) | 522 (53.8%) |
|  | no, but facility has capability | 1 (3.2%) | 50 (6.1%) | 5 (4.3%) | 56 (5.8%) |
|  | no/unknown | 7 (22.6%) | 351 (42.5%) | 35 (30.4%) | 393 (40.5%) |
| Caesarean section | yes | 22 (71.0%) | 208 (25.2%) | 59 (51.8%) | 289 (29.8%) |
|  | not in last 12 months, but has capability | 0 (0.0%) | 22 (2.7%) | 5 (4.4%) | 27 (2.8%) |
|  | no capability | 9 (29.0%) | 591 (71.6%) | 50 (43.9%) | 650 (67.0%) |
|  | only post-operative care | 0 (0.0%) | 4 (0.5%) | 0 (0.0%) | 4 (0.4%) |
| Blood transfusion | yes | 27 (87.1%) | 492 (59.6%) | 89 (77.4%) | 608 (62.6%) |
|  | no, but facility has capability | 1 (3.2%) | 50 (6.1%) | 9 (7.8%) | 60 (6.2%) |
|  | no/unknown | 3 (9.7%) | 283 (34.3%) | 17 (14.8%) | 303 (31.2%) |
| BEmONC (7 functions performed) | | 7 (22.6%) | 179 (21.7%) | 21 (18.3%) | 207 (21.3%) |
| BEmONC-1 (6 functions performed ) | | 17 (54.8%) | 325 (39.4%) | 57 (49.6%) | 399 (41.1%) |
| CEmONC (9 functions performed) | | 5 (16.1%) | 56 (6.8%) | 12 (10.4%) | 73 (7.5%) |
| CEmONC-1 (8 functions performed ) | | 13 (41.9%) | 128 (15.5%) | 37 (32.2%) | 178 (18.3%) |

**Supplementary Table 4.** **Reported performance of emergency obstetric care functions (in the previous 12 months), by level (among facilities providing childbirth care n= 971)**

| Variable | Category | Primary | Secondary | Tertiary | All facilities |
| --- | --- | --- | --- | --- | --- |
| N |  | 853 | 116 | 2 | 971 |
| Administration of IV antibiotics | yes | 808 (94.7%) | 112 (96.6%) | 2 (100.0%) | 922 (95.0%) |
|  | no, but facility has capability | 24 (2.8%) | 4 (3.4%) | 0 (0.0%) | 28 (2.9%) |
|  | no/unknown | 21 (2.5%) | 0 (0.0%) | 0 (0.0%) | 21 (2.2%) |
| Administration of IV/IM oxytocics | yes | 792 (92.8%) | 111 (95.7%) | 2 (100.0%) | 905 (93.2%) |
|  | no, but facility has capability | 40 (4.7%) | 3 (2.6%) | 0 (0.0%) | 43 (4.4%) |
|  | no/unknown | 21 (2.5%) | 2 (1.7%) | 0 (0.0%) | 23 (2.4%) |
| Administration of parenteral magnesium sulphate for eclampsia | yes | 626 (73.4%) | 96 (82.8%) | 2 (100.0%) | 724 (74.6%) |
|  | no, but facility has capability | 130 (15.2%) | 15 (12.9%) | 0 (0.0%) | 145 (14.9%) |
|  | no/unknown | 97 (11.4%) | 5 (4.3%) | 0 (0.0%) | 102 (10.5%) |
| Assisted vaginal birth | yes | 283 (33.2%) | 41 (35.3%) | 0 (0.0%) | 324 (33.4%) |
|  | no, but facility has capability | 68 (8.0%) | 18 (15.5%) | 0 (0.0%) | 86 (8.9%) |
|  | no/unknown | 502 (58.9%) | 57 (49.1%) | 2 (100.0%) | 561 (57.8%) |
| Manual extraction of placenta | yes | 738 (86.5%) | 102 (87.9%) | 2 (100.0%) | 842 (86.7%) |
|  | no, but facility has capability | 84 (9.8%) | 12 (10.3%) | 0 (0.0%) | 96 (9.9%) |
|  | no/unknown | 31 (3.6%) | 2 (1.7%) | 0 (0.0%) | 33 (3.4%) |
| Removal of retained products of conception | yes | 780 (91.4%) | 105 (90.5%) | 2 (100.0%) | 887 (91.3%) |
|  | no, but facility has capability | 48 (5.6%) | 10 (8.6%) | 0 (0.0%) | 58 (6.0%) |
|  | no/unknown | 25 (2.9%) | 1 (0.9%) | 0 (0.0%) | 26 (2.7%) |
| Neonatal resuscitation with bag and mask | yes | 428 (50.2%) | 92 (79.3%) | 2 (100.0%) | 522 (53.8%) |
|  | no, but facility has capability | 51 (6.0%) | 5 (4.3%) | 0 (0.0%) | 56 (5.8%) |
|  | no/unknown | 374 (43.8%) | 19 (16.4%) | 0 (0.0%) | 393 (40.5%) |
| Caesarean section | yes | 202 (23.7%) | 85 (73.3%) | 2 (100.0%) | 289 (29.8%) |
|  | not in last 12 months, but has capability | 23 (2.7%) | 4 (3.4%) | 0 (0.0%) | 27 (2.8%) |
|  | no capability | 624 (73.2%) | 26 (22.4%) | 0 (0.0%) | 650 (67.0%) |
|  | only post-operative care | 3 (0.4%) | 1 (0.9%) | 0 (0.0%) | 4 (0.4%) |
| Blood transfusion | yes | 509 (59.7%) | 97 (83.6%) | 2 (100.0%) | 608 (62.6%) |
|  | no, but facility has capability | 54 (6.3%) | 6 (5.2%) | 0 (0.0%) | 60 (6.2%) |
|  | no/unknown | 290 (34.0%) | 13 (11.2%) | 0 (0.0%) | 303 (31.2%) |
| BEmONC (7 functions performed) |  | 176 (20.6%) | 31 (26.7%) | 0 (0.0%) | 207 (21.3%) |
| BEmONC-1 (6 functions performed ) |  | 328 (38.5%) | 69 (59.5%) | 2 (100.0%) | 399 (41.1%) |
| CEmONC (9 functions performed) |  | 53 (6.2%) | 20 (17.2%) | 0 (0.0%) | 73 (7.5%) |
| CEmONC-1 (8 functions performed ) |  | 121 (14.2%) | 55 (47.4%) | 2 (100.0%) | 178 (18.3%) |

**Supplementary Figure 1. A: Emergency obstetric care functions by facility sector. B – Emergency care functions by facility level. N=971 facilities providing childbirth care in Lubumbashi.**

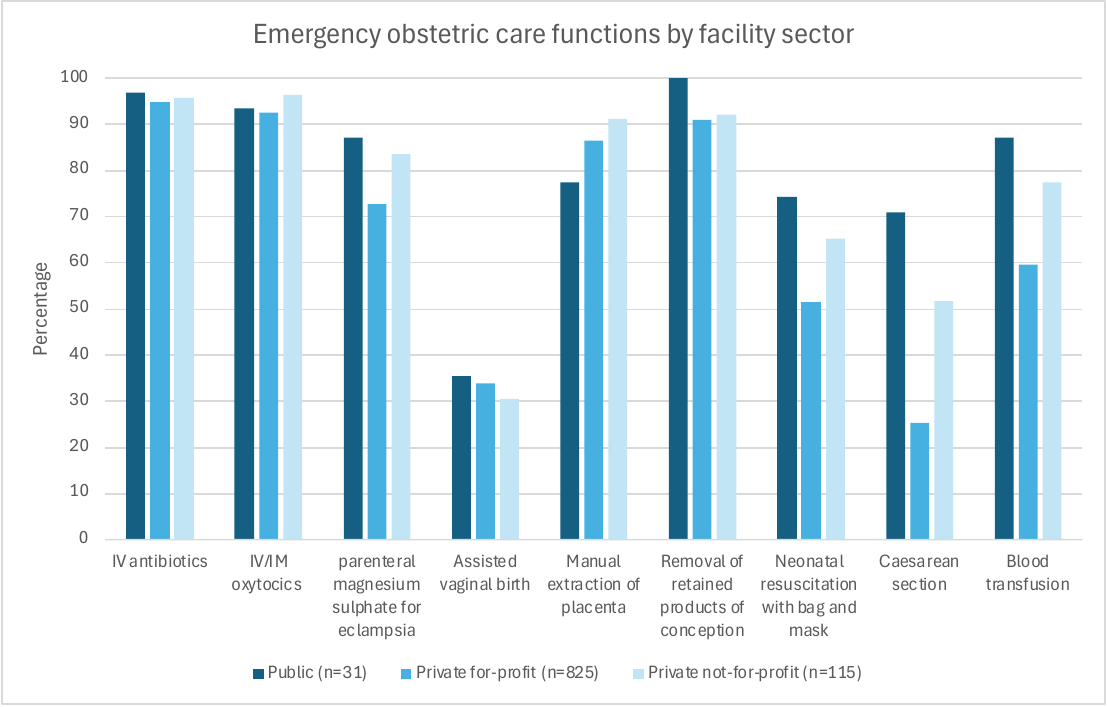

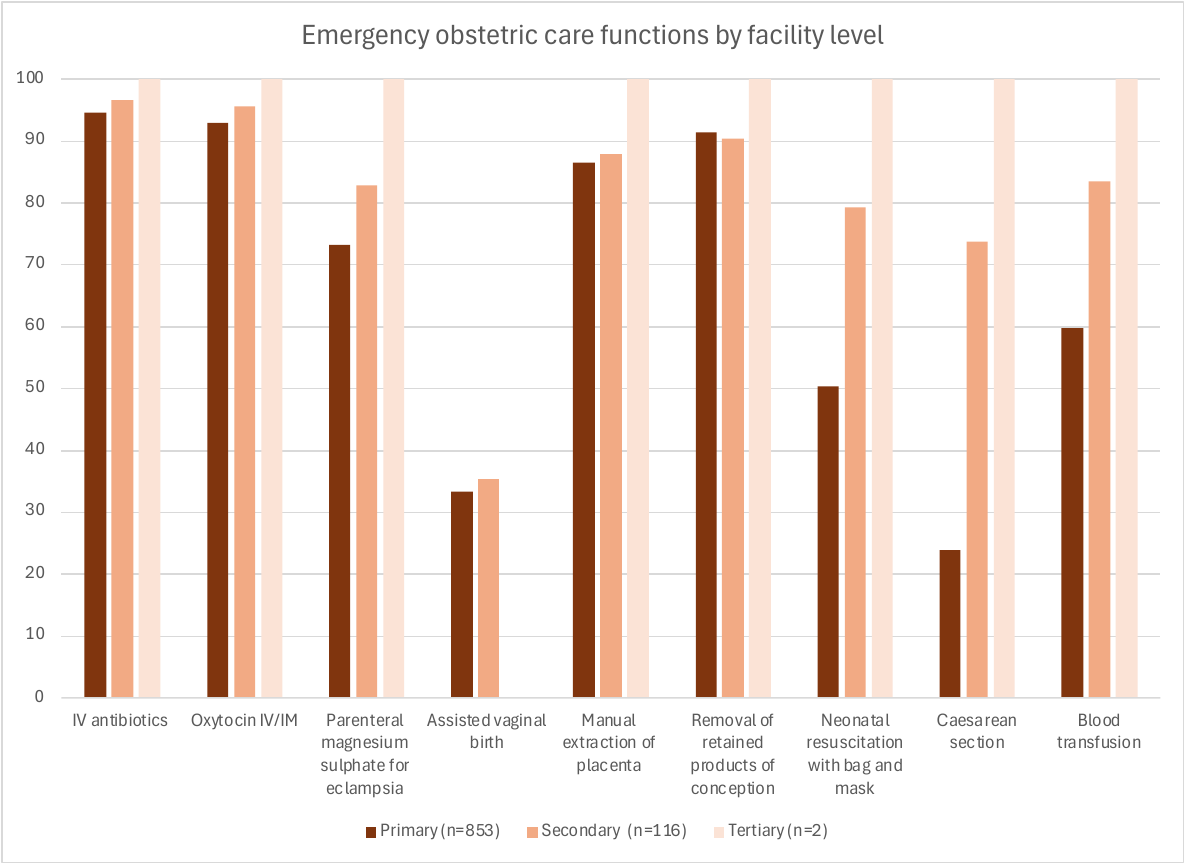

**Supplementary Table 5.** Number and percentage of facilities by reported birth volume category, by health zone, Lubumbashi (n=971)

| Health zone | <10 per month (<120/year) | 10-30 per month (120-360/year) | >30 per month (>360/year) | Does not know | Total |
| --- | --- | --- | --- | --- | --- |
| Kamalondo | 5 (50.0%) | 4 (40.0%) | 1 (10.0%) | 0 (0.0%) | 10 |
| Kampemba | 108 (56.8%) | 75 (39.5%) | 7 (3.7%) | 0 (0.0%) | 190 |
| Katuba | 14 (38.9%) | 16 (44.4%) | 5 (13.9%) | 1 (2.7%) | 36 |
| Kenya | 35 (44.9%) | 36 (45.3%) | 7 (9.0%) | 0 (0.0%) | 78 |
| Kisanga | 31 (33.0%) | 56 (59.6%) | 7 (7.5%) | 0 (0.0%) | 94 |
| Kowe | 1 (20%) | 4 (80%) | 0 (0.0%) | 0 (0.0%) | 5 |
| Lubumbashi | 85 (49.4%) | 77 (44.8%) | 9 (5.2%) | 1 (0.6%) | 172 |
| Mumbunda | 73 (56.2%) | 48 (36.9%) | 9 (6.9%) | 0 (0.0%) | 130 |
| Ruashi | 86 (47.1%) | 81 (44.8%) | 14 (7.7%) | 0 (0.0%) | 181 |
| Tshamilemba | 28 (39.4%) | 36 (50.7%) | 7 (9.9%) | 0 (0.0%) | 71 |
| Vangu | 0 (0.0%) | 3 (75.0%) | 1 (25.0%) | 0 (0.0%) | 4 |
| **Lubumbashi City (total)** | **466 (48.0%)** | **436 (44.9%)** | **67 (6.9%)** | **2 (0.2%)** | **971** |

**Supplementary Table 6.** **Summary of missingness of select indicators extracted from Maternity Ward registers (133 facilities) during 12-month period (November 2022-October 2023), including median and range across facilities**

|  | N  (facilities) | Facilities with at least one missing month (%) | Facilities with at six or more (out of 12) missing months (%) | Facilities with all (out of 12) missing months (%) | Median | Range |
| --- | --- | --- | --- | --- | --- | --- |
| Births | 133 | 53 (39.8%) | 10 (7.5%) | 1 (0.8%) | 298 | 8-904 |
| C-sections | 102 | 60 (58.8%) | 27 (26.5%) | 0 | 11 | 1-199 |
| Live births | 133 | 62 (46.6%) | 23 (17.3%) | 11 (8.3%) | 238 | 0-891 |
| Stillbirths | 133 | 102 (76.7%) | 70 (52.6%) | 28 (21.1%) | 2 | 0-104 |
| Eclampsia | 133 | 104 (78.2%) | 69 (51.9%) | 36 (27.1%) | 1 | 0-82 |
| Antepartum hemorrhage | 133 | 106 (79.7%) | 74 (55.6%) | 38 (28.6%) | 1 | 0-73 |
